# Gut dysbiosis and metabolic disruption distinguish continence outcomes after anorectal malformation repair

**DOI:** 10.1101/2025.10.26.25338825

**Authors:** Bhavin Parekh, Aditi Kaloni, Rakesh S Joshi, Jaishri Ramji, Swayamprabha Samantaray, Anupama Modi

**Affiliations:** School of Applied Sciences and Technology, Gujarat Technological University, Ahmedabad, Gujarat, India; Department of Pediatric Surgery, Civil Hospital, Ahmedabad, Gujarat, India; Department of Validation of Indic Knowledge through Advanced Research, Gujarat University, Ahmedabad, Gujarat, India

**Author notes:** **Corresponding authors: Dr. Bhavin Parekh,** Research Fellow, Department of Validation of Indic Knowledge through Advanced Research, Gujarat University, Gujarat - 380009., **Dr. Rakesh S Joshi,** Head of Department, Department of Pediatric Surgery, Civil Hospital, Gujarat – 380016, **Dr. Anupama Modi,** Assistant Professor, School of Applied Sciences and Technology, Gujarat Technological University, Gujarat - 382424.

**Keywords:** Faecal incontinence, Anorectal malformation, Gut microbiome, Metabolomics, Enteric nervous system

## Abstract

Faecal incontinence (FI) is a frequent and debilitating sequela of surgical repair of anorectal malformations (ARMs). The rectoanal inhibitory reflex (RAIR), an enteric nervous system (ENS)-mediated circuit essential for continence, is often absent or impaired in these children post-surgically. However, the biological cascade linking surgery to this reflex failure remains unknown. To identify the mechanistic pathways underlying post-surgical FI, we performed a multi-omics case-control study of 31 post-ARM children (12 incontinent, 20 continent), integrating stool 16S rRNA sequencing with untargeted serum metabolomics by liquid chromatography-mass spectrometry (LC-MS). Incontinent children exhibited greater microbial richness yet distinct communities, marked by depletion of butyrate-producing *Faecalibacterium* and expansion of mucin-degrading *Ruminococcus* along with Proteobacteria. These ecological shifts coincided with signatures of impaired fatty acid oxidation, and elevated levels of neurotoxic and inflammatory compounds such as trimethylamine and kynurenines. These findings suggest that surgical trauma destabilizes microbial and metabolic homeostasis, compromising ENS circuits. These results raise the possibility that microbial and metabolic restoration could restore faecal continence after surgical repair.

**Graphical Abstract:** 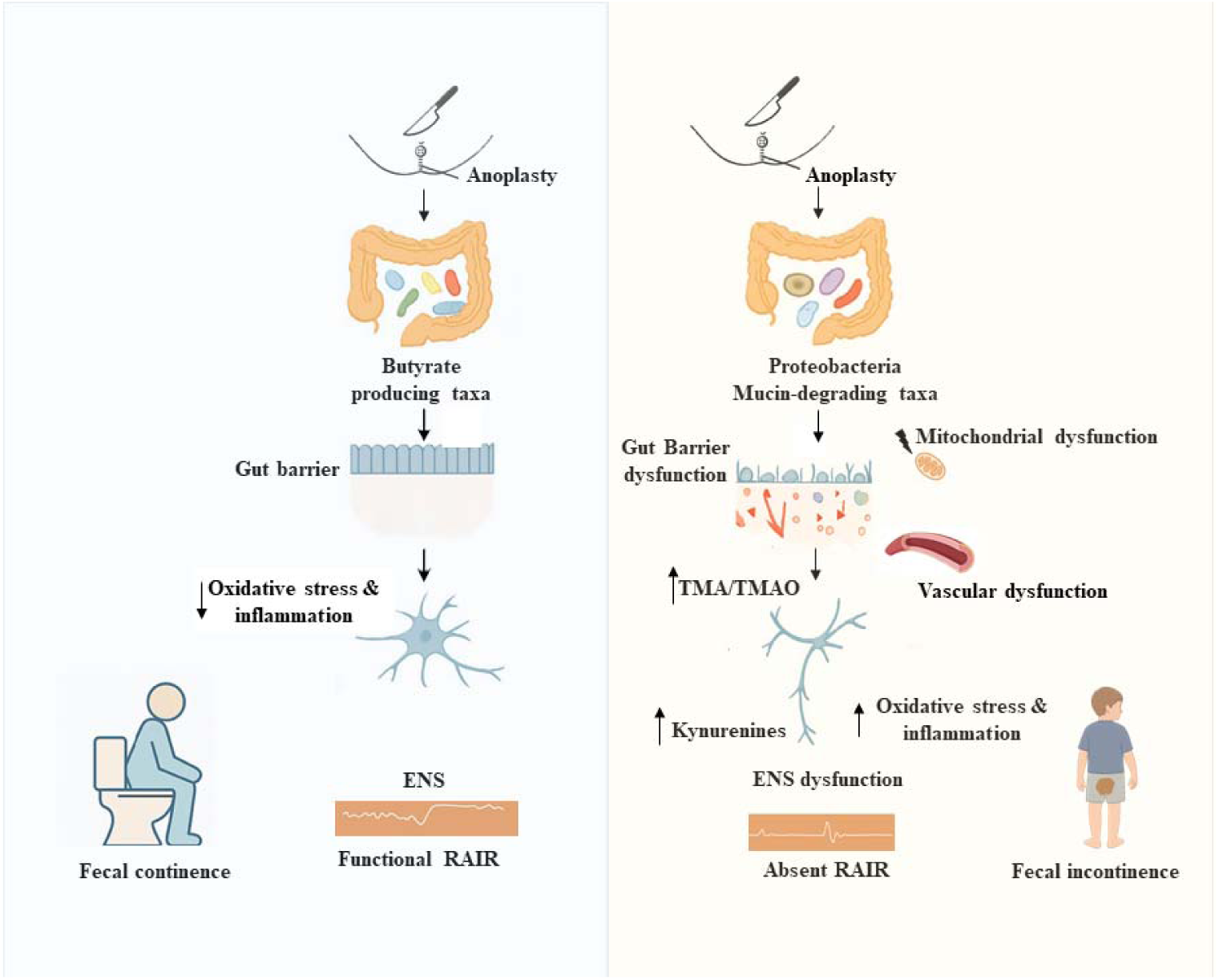

## Introduction

Anorectal malformations (ARMs) occur in roughly 1 in 5,000 newborns and demand early surgical correction to restore continuity of the anorectal canal^1,2^. Modern techniques like posterior sagittal anorectoplasty and laparoscopy-assisted approaches can precisely reconstruct the anorectum^1,2^. It is therefore a paradox that this structural success so often begets a functional failure—faecal incontinence (FI)^2–5^. This failure is rooted in the loss of the rectoanal inhibitory reflex (RAIR), an enteric nervous system (ENS) circuit essential for continence^6^. The RAIR triggers internal anal sphincter relaxation during rectal distension, permitting sampling of rectal contents (i.e., gas and stool) and coordinating involuntary internal anal sphincter activity with voluntary external anal sphincter control for continence□’□. Disruption of this reflex impairs rectal emptying, leads to megarectum (constipation), and eventually results in soiling due to overflow incontinence^6^. How an anatomically precise operation disrupts this neural reflex remains an enigma in paediatric surgery.

We contend that the answer lies not at the surgeon’s scalpel, but within the gut microbiota. Surgery is an ecological shock to this microbial ecosystem, disrupting the symbiotic equilibrium that sustains gastrointestinal health and neural function^7^. The combined insults of surgical trauma, tissue ischemia, and antibiotic use are known to destabilize the gut microbiota, depleting butyrate-producing commensals that support neuronal health while promoting colonization of pathobionts^8–12^. Notably, gut dysbiosis in other contexts has been associated with epithelial barrier dysfunction and pro-inflammatory conditions that affect ENS function ^11–15^. Indeed, emerging evidence links microbiome composition to ENS function, development, and plasticity¹³□¹□. Whether this post-surgical dysbiosis underlies continence failure, however, has not been explored. This study was therefore designed to investigate this ecological hypothesis. Using multi-omics, we surveyed the microbial and metabolic profiles of children with and without post-surgical FI. We reveal that children with FI harbor distinct microbial and metabolic signatures. Our results indicate that post-surgical FI may constitute a remediable perturbation of the gut–neural axis, rather than an unavoidable surgical shortcoming.

## Method

### Study Design and Participants

We conducted a single-centre, cross-sectional case–control study at Civil Hospital, Ahmedabad, India, between December 2023 and October 2024. Children with anorectal malformation repair completed ≥3 months prior were recruited during routine follow-up. Participants were stratified into faecal incontinence (cases, *n* = 12) or continent (controls, *n* = 20) cohorts based on Krickenbeck and Rintala scoring systems (Figure 1). Faecal incontinence was defined as involuntary passage of liquid or solid stool^16^.

**Figure 1.**
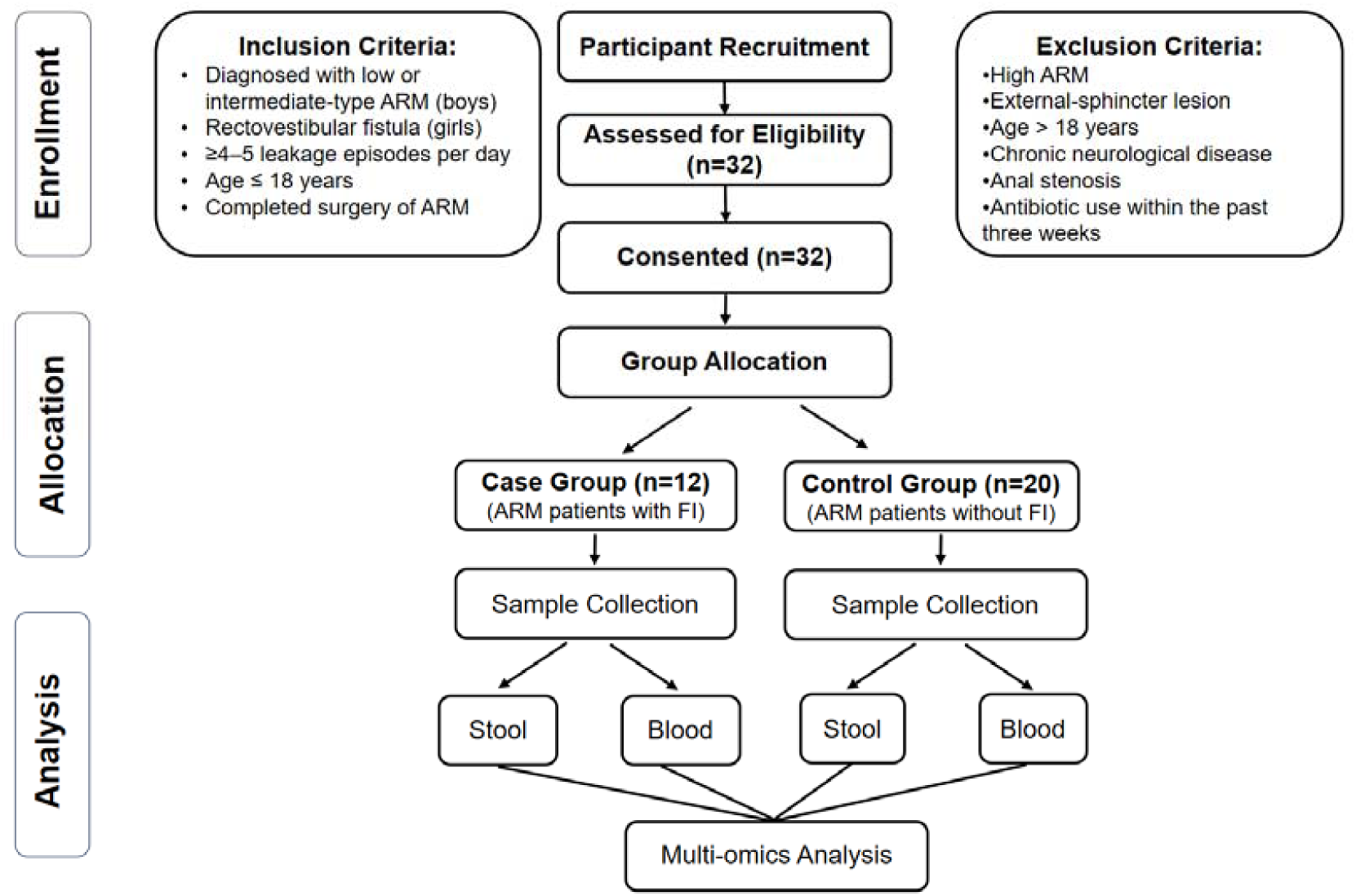
Participant enrollment, grouping, and sample collection workflow. Children with anorectal malformation (ARM) were screened according to predefined inclusion and exclusion criteria, and 32 eligible participants were enrolled. Participants were stratified into two groups: children with faecal incontinence (FI; n = 12) and continent controls (n = 20). Stool and blood samples were collected from each participant for downstream multi-omics analyses, including microbiome and metabolome profiling.

### Inclusion and Exclusion Criteria

Inclusion criteria were low- or intermediate-type ARM in boys, rectovestibular fistula in girls, and ≥4 leakage episodes daily. Exclusion criteria included high ARM, incomplete repair, external sphincter injury, active infection, age >18 years, chronic neurological disease, anal stenosis, or antibiotic use within three weeks of enrolment (Figure 1).

### Ethical Approval

The protocol was approved by the Civil Hospital Institutional Ethics Committee (EC/Approval/107/2023/04/12/2023). Written informed consent was obtained from parents or legal guardians in Hindi, Gujarati or English. All procedures conformed to National Accreditation Board for Hospitals and Healthcare Providers (NABH) guidelines.

### Clinical and Demographic Data Collection

Demographic and clinical variables were recorded to control for confounding: age, sex, diet (vegetarian/non-vegetarian), delivery mode, pregnancy complications, miscarriage history, gestational age, stool consistency, toilet-training status, and maternal blood pressure.

### Sample Collection and Processing

Fresh stool was passed into sterile containers, immediately placed on dry ice and stored at −80 °C. Venous blood was collected into serum-separator tubes, incubated 60 min at room temperature, centrifuged (1300 *g* for 5 min) and the serum aliquoted (0.5 mL) and frozen at −80 °C. The samples were processed at the Gujarat Biotechnology Research Center (GBRC) and Gujarat Technological University (GTU).

### DNA Extraction and 16S rRNA Gene Sequencing

Microbial DNA was extracted from 250 mg stool using the HiPurA Stool DNA Purification Kit (HiMedia, India). DNA quantity, purity (NanoDrop spectrophotometer, Qubit fluorometer), and integrity (1% agarose gel) were assessed; only samples with A_260/280_ ratios of 1.8–2.0 and concentrations ≥20 ng μL□¹ were retained. The V3–V4 region of the 16S rRNA gene was amplified using primers 341F and 785R with Illumina overhangs in 25 μL reactions containing 2× KAPA HiFi HotStart ReadyMix (95 °C for 3 min; 25 cycles of 95 °C for 30 s, 55 °C for 30 s, 72 °C for 30 s; 72 °C for 5 min). Amplicons were purified with AMPure XP beads (Beckman Coulter), indexed using Nextera XT adapters, quantified by Qubit, and pooled. The library was sequenced on an Illumina MiSeq platform (2 × 250 bp paired-end reads, v3 chemistry) with 5% PhiX spike-in.

### Microbiome Data Processing and Analysis

Raw sequences were processed in QIIME2 (v2024.2). Reads were trimmed to 250 bp, low-quality and chimeric sequences were removed with VSEARCH, and amplicon sequence variants (ASVs) were inferred using Deblur. After denoising and chimera removal, taxonomy was assigned against the GreenGenes database (v13_8, 99% similarity), and ASVs with <10 total reads were excluded. α-diversity was assessed using Shannon and Chao1 indices, and β-diversity using Bray–Curtis, Jaccard, weighted, and unweighted UniFrac metrics. Rarefaction depth was set to 1000 reads per sample, and group differences in β-diversity were tested by PERMANOVA (999 permutations). Taxonomic composition was profiled at phylum, family, and genus levels.

### Untargeted Serum Metabolomics Analysis

Serum metabolites were extracted by protein precipitation: 100 μL serum was mixed with 900 μL ice-cold acetonitrile, vortexed for 30 s, and centrifuged at 12,500 rpm for 15 min at 4 °C. Supernatants were transferred to autosampler vials for analysis. Metabolite profiling was performed on an Agilent 6545XT Advanced Bio LC-QTOF mass spectrometer in positive electrospray ionization mode (50–1200 m/z). Chromatographic separation used a C18 column (2.1 × 100 mm, 1.7 μm) at 35 °C with 0.200 mL/min flow rate. Mobile phases comprised 0.1% formic acid in water (A) and acetonitrile (B), with 25-min gradient elution (1–95% B). Pooled quality control (QC) samples were injected after each batch to monitor system stability; blank samples assessed background interference. Metabolites with coefficient of variation >30% across QC samples were excluded.

Raw data files were converted to .mzML format using ProteoWizard and processed in MS-DIAL for feature detection, spectral deconvolution, metabolite identification, and peak alignment. Peak detection was performed with a minimum height of 1000 amplitude and mass slice width of 0.05 Da. MS2Dec deconvolution used a sigma window value of 0.5 and MS/MS abundance cutoff of 10 amplitude. Metabolite identification was conducted with a mass tolerance of 0.005 Da and identification score cutoff of 80%. Alignment was performed with retention time tolerance of 0.05 min and MS1 tolerance of 0.005 Da. Peak picking used minimum height of 20,000 counts and minimum width of 10 scans. Peak alignment employed retention time tolerance of 0.5 min and mass tolerance of 0.0015 Da. Data were normalized, log-transformed, and Pareto-scaled using MetaboAnalyst v6.0 with abundance filtering and quantile normalization.

### Multivariate and Univariate Statistical Analysis of Metabolomics

Principal Component Analysis (PCA) was used to examine group-level variance. Supervised classification was performed using Orthogonal Partial Least Squares Discriminant Analysis (OPLS-DA), validated through permutation testing. VIP scores quantified each metabolite’s contribution to group discrimination. Univariate tests included fold change, independent t-tests, and Mann–Whitney U tests. Differentially expressed metabolites (DEMs) were selected using thresholds: p < 0.05, fold change >1.5, and VIP >1.0. Overlapping hits across multivariate and univariate analyses were visualized using Venn diagrams. Thirty DEMs were shortlisted.

### Pathway and Disease Association Analysis

MetaboAnalyst’s enrichment module revealed 45 significant pathways (FDR < 0.05), notably involving amino acid metabolism, FMO-mediated oxidation, and trimethylamine-N-oxide processing. Disease enrichment via the Human Metabolome Database connected the 30 DEMs to 54 traits, including preeclampsia, diabetes mellitus type 2, and uremia.

### Integration and Correlation Analysis

Microbiome–metabolome integration was conducted using Spearman’s rank correlation. Correlation matrices were FDR-adjusted, and associations with *p* < 0.05 were considered significant. Significant correlations were visualized as heatmaps to highlight biologically relevant patterns.

### Statistical Analysis

Descriptive statistics included mean ± SD for normally distributed variables, and median (IQR) for skewed data. Shapiro–Wilk test evaluated normality. Group differences were assessed using t-tests, Mann–Whitney U tests, or Fisher’s Exact Test (for categorical data). β-diversity group differences were tested via PERMANOVA. All visualizations—heatmaps, volcano plots, PCA plots, and taxonomic trees—were generated using ggplot2 and pheatmap packages in R (v4.3.1). Statistical significance was set at p < 0.05 throughout.

## Results

### Participant Characteristics

Thirty-two children with repaired anorectal malformations were enrolled—12 cases and 19 controls. Age (median 5 ± 2.7 yr vs 5 ± 4 yr; Mann–Whitney *p* = 0.56) and sex distribution (males 42% vs 55%; χ² *p* = 0.29) were comparable between groups; however, diet and delivery mode differed (Table 1). Vegetarian diet predominated in cases (92% vs 50%; χ² *p* = 0.023), while caesarean section was more frequent (50% vs 15%; χ² *p* = 0.049) and vaginal delivery less common (50% vs 85%) (Table 2). Pregnancy complications, maternal miscarriage, prematurity, gestational hypertension, stool consistency, and toilet-training status showed no significant differences (Table 3). Vegetarian diet and caesarean delivery were the only variables enriched in cases, whereas other demographic, perinatal, and clinical features were similar.

**Table 1.**
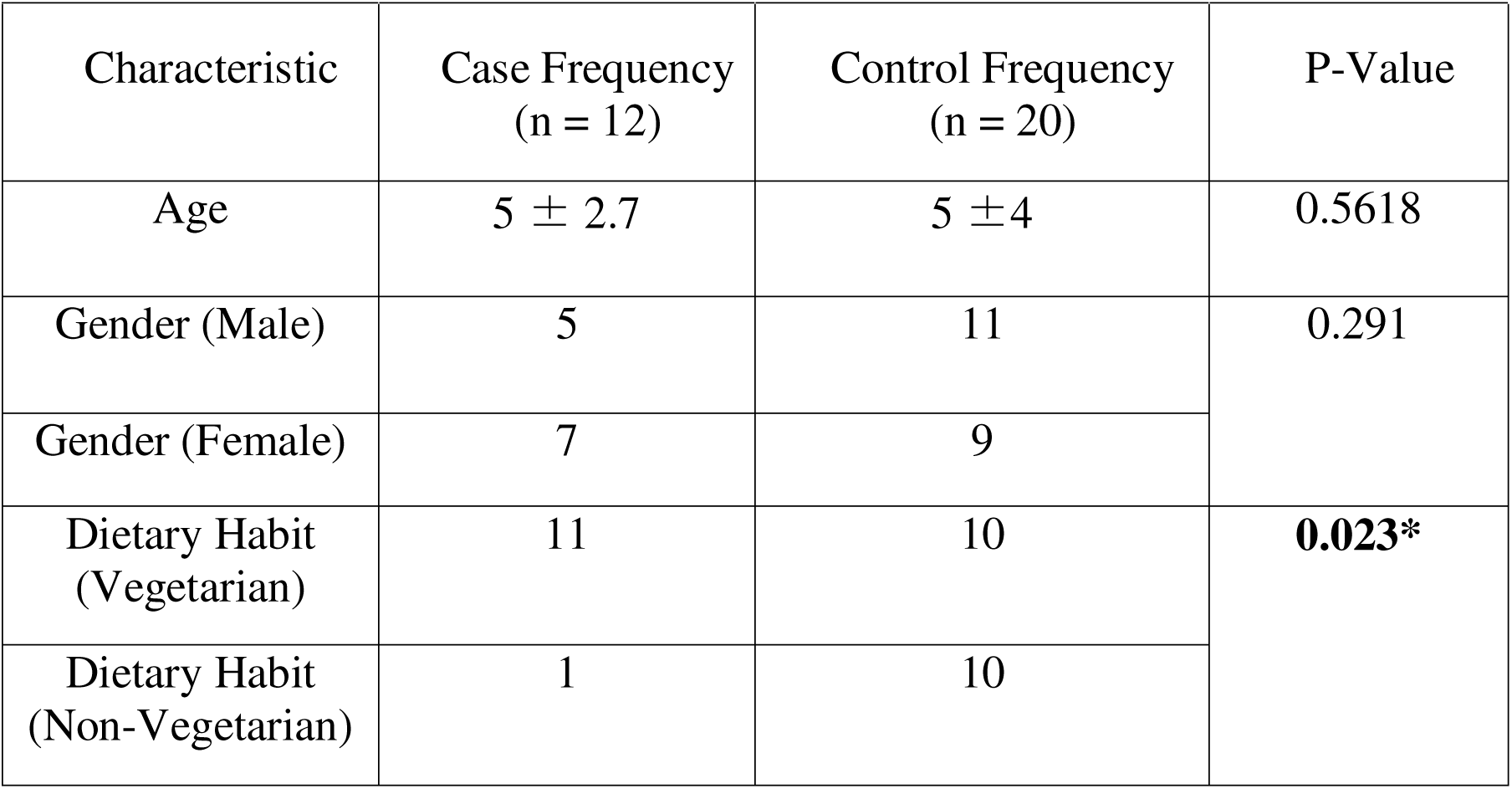
Clinical characteristics of children with FI and controls. Comparison of demographic and clinical parameters between children with FI (n = 12) and continent controls (n = 20). Data are presented as n (%) unless otherwise indicated. Group differences were evaluated using two-sided χ² tests or Fisher’s exact tests, as appropriate, and corresponding p-values are reported.

**Table 2.**
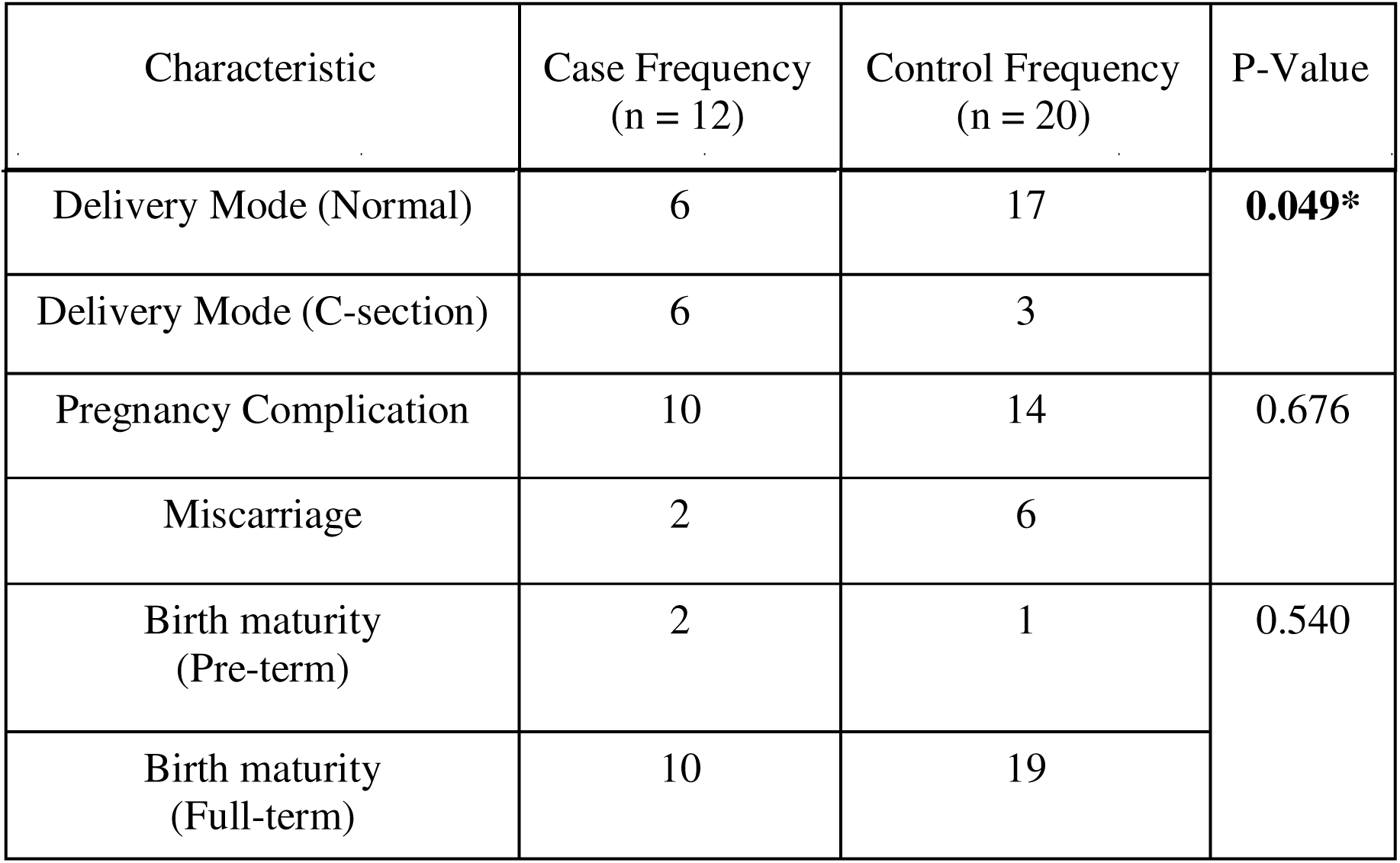
Perinatal and obstetric characteristics of children with FI and controls. Comparison of perinatal and maternal parameters between children with FI (n = 12) and continent controls (n = 20). Data are presented as n (%) unless otherwise indicated. Group differences were evaluated using two-sided χ² tests or Fisher’s exact tests, as appropriate, and corresponding p-values are reported.

**Table 3.** Bowel-function and maternal parameters in children with FI and controls. Comparison of stool consistency, toilet-training status, and maternal gestational blood pressure between children with FI (n = 12) and continent controls (n = 20). Data are expressed as n (%) unless otherwise indicated. Statistical significance was determined using two-sided χ² or Fisher’s exact tests, with p-values reported..

| Characteristic | Case Frequency<br>(n = 12) | Control<br>Frequency<br>(n = 20) | P-Value |
| --- | --- | --- | --- |
| Stool Consistency<br>(Normal) | 11 | 16 | 0.123 |
| Stool Consistency<br>(Hard) | 0 | 4 | 0.054 |
| Stool Consistency<br>(Liquid) | 1 | 0 | 0.846 |
| Toilet Trained<br>(No) | 1 | 0 | 0.375 |
| Toilet Trained<br>(Yes) | 11 | 20 |  |
| Gestational BP<br>(Normal) | 9 | 15 | 1.00 |
| Gestational BP<br>(High) | 3 | 5 |  |

### Microbiome Analysis

Gut-microbiome profiling revealed consistent divergence between cases and controls. Cases showed numerically higher α-diversity richness (Chao1 median ≈ 195 vs 120; Mann–Whitney *p* = 0.527) (Figure 2a) and evenness (Shannon median ≈ 3.2 vs 2.9; *p* = 0.104) (Figure 2b) compared with controls, although neither difference reached statistical significance. β-diversity principal-coordinate analyses demonstrated distinct clustering: Bray–Curtis and Jaccard plots (Figure 3a & b) showed partially overlapping yet distinct clusters, while weighted and unweighted UniFrac metrics (Figure 3c & d) reinforced this separation—the weighted metric highlighting abundance-driven phylogenetic divergence. The gut microbiome of cases was not only richer and more even but also compositionally distinct, patterns potentially relevant to postoperative bowel dysfunction.

**Figure 2.**
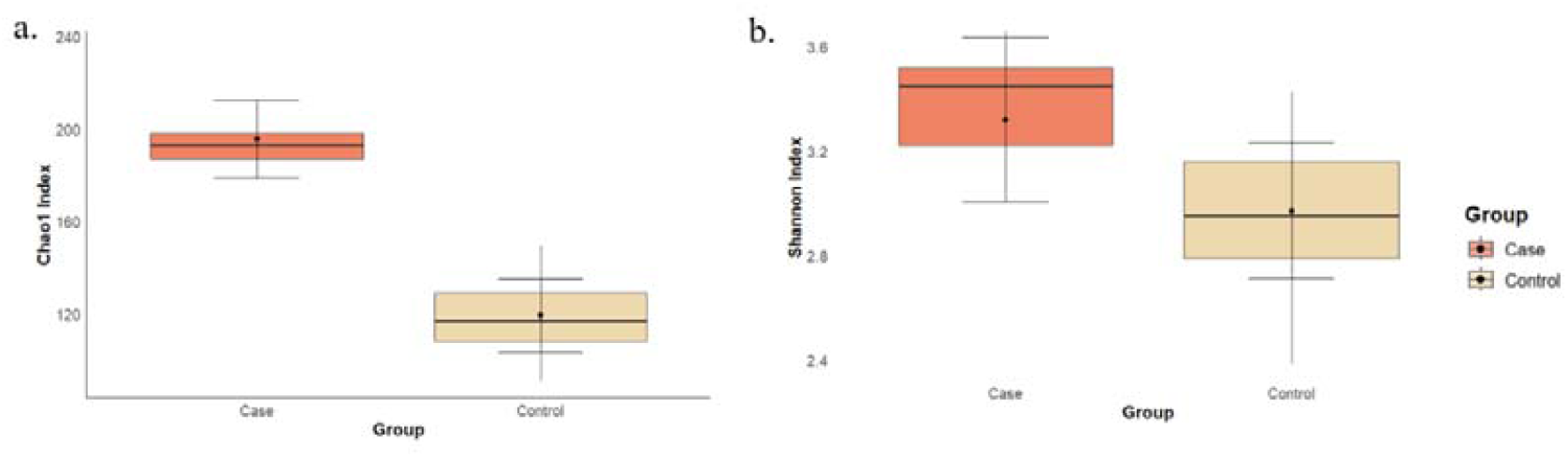
α-diversity metrics comparing faecal microbiota of children with FI and controls. (a) Chao1 index representing species richness. (b) Shannon index reflecting both richness and evenness. Children with FI (n = 12) showed numerically higher microbial richness (Chao1 median ≈ 195 vs 120) and evenness (Shannon median ≈ 3.2 vs 2.9) compared with continent controls (n = 20), though differences were not statistically significant (Mann–Whitney: Chao1 p = 0.527, Shannon p = 0.104). Box plots display the median (centre line), interquartile range (box), and minimum– maximum whiskers; black dots indicate mean values. Colours denote groups: FI (red) and controls (beige).

**Figure 3.**
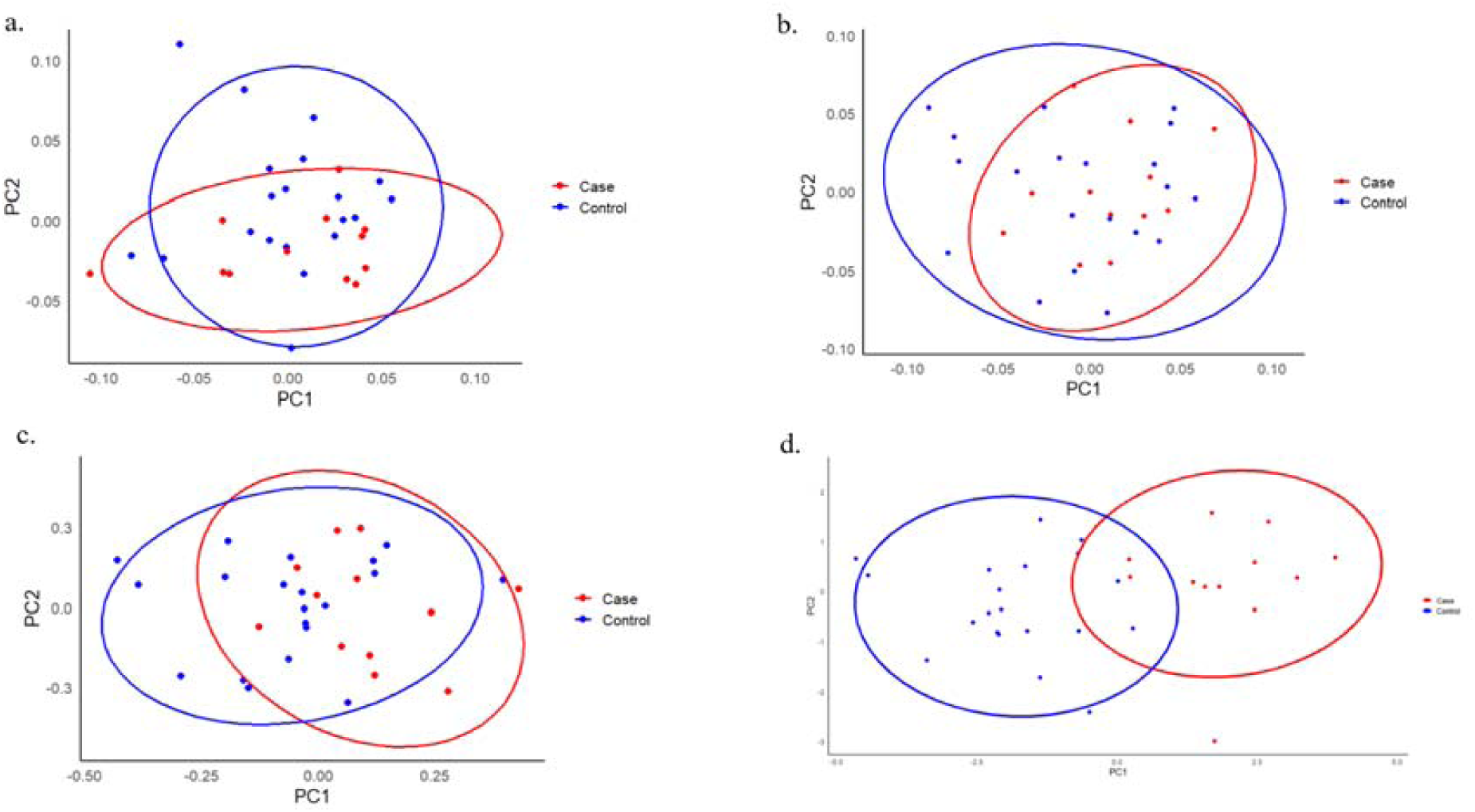
β-diversity of faecal microbial communities in children with FI and controls. Principal coordinate analysis (PCoA) illustrating differences in microbial community composition between FI (n = 12) and control (n = 20) groups. Plots show ordinations based on (a) Bray–Curtis, (b) Jaccard, (c) weighted UniFrac, and (d) unweighted UniFrac distance metrics. Ellipses represent 95% confidence intervals for each group. Group separation was evaluated using PERMANOVA.

### Taxonomic Composition Analysis

At the phylum level, cases and controls showed distinct profiles: Firmicutes dominated both groups but were slightly higher in cases (59.31%) than controls (56.7%); Bacteroidetes were reduced in cases (15.06% vs 23.48%), resulting in an elevated Firmicutes/Bacteroidetes ratio (3.94 vs 2.41) (Supplementary Figure 1); Actinobacteria increased modestly (16.8% vs 13.6%); and Proteobacteria— often associated with inflammation—were elevated (9.3% vs 6.6%) (Figure 4a). At the family level, Ruminococcaceae were depleted in cases (51.2% vs 79.3%), whereas Lachnospiraceae were higher (49.8% vs 36.2%); Sphingobacteriaceae and Coriobacteriaceae were similar across groups (6.4% vs 6.8% and 2.9% vs 3.2%, respectively), and Pasteurellaceae (Gammaproteobacteria) were increased (1.2% vs 0.4%) (Figure 3b). At the genus level, *Faecalibacterium* was depleted in cases (66% vs 80%), whereas *Roseburia* (32% vs 5%) and *Ruminococcus* (20% vs 2%) were enriched; minor shifts included slightly higher Clostridium spp. in controls (6% vs 3%), with *Butyricicoccus* comparable between groups (2% vs 3%) (Figure 4c).

**Figure 4.**
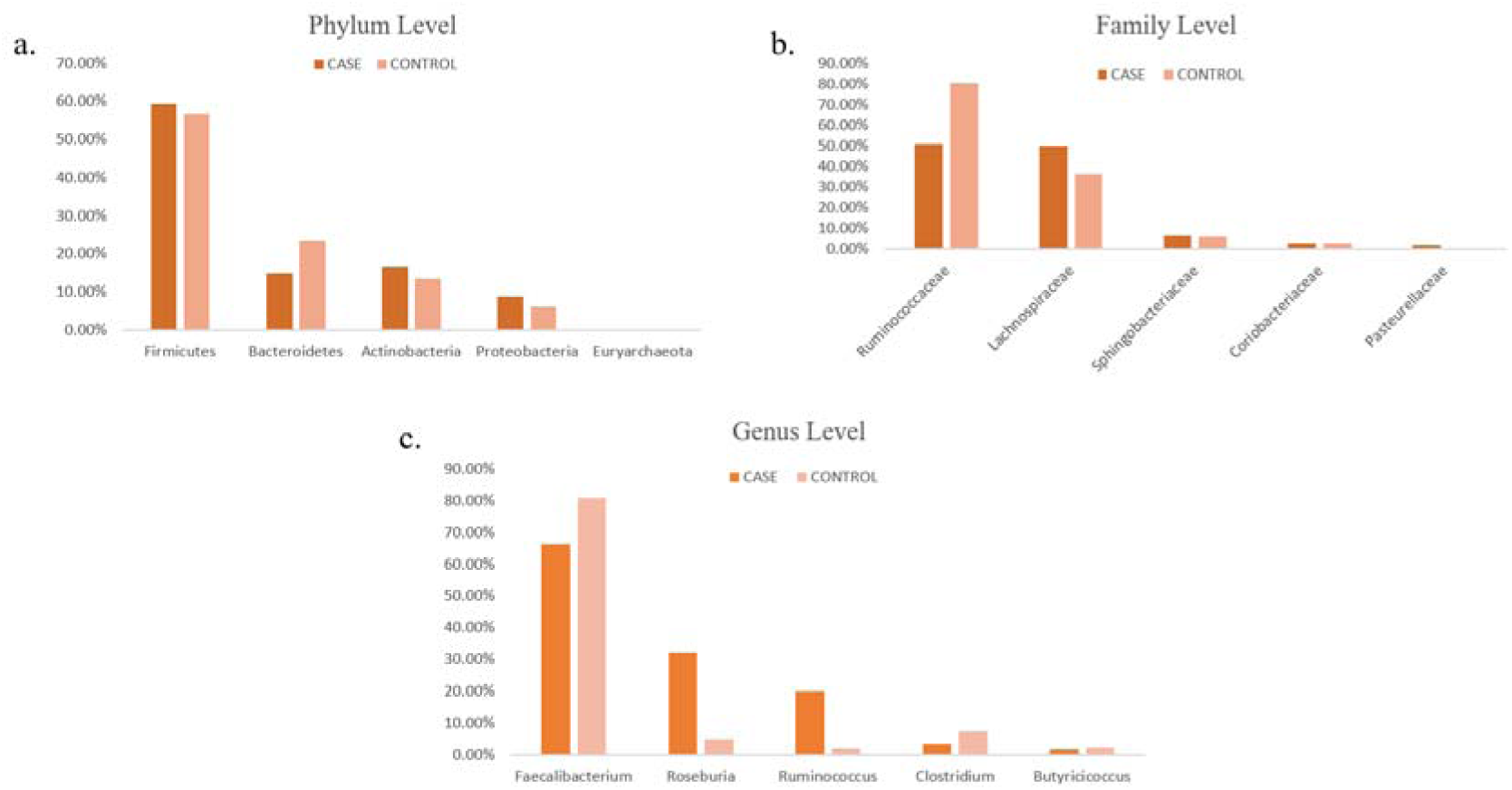
Taxonomic composition of the gut microbiota in children with faecal FI and controls. Relative abundance of bacterial taxa at (a) phylum, (b) family, and (c) genus levels in children with FI (n = 12) and continent controls (n = 20). Bar plots display mean re ative abundances with error bars representing standard error. Statistical comparisons were performed using the two-sided Mann–Whitney U test. Significance levels are indicated as follows: ns, not significant; *p□*<*□*0.05.

### Serum Metabolomics Analysis

Targeted profiling identified 25 differentially abundant serum metabolites that clearly separated cases from controls (Figure 5a). PLS-DA highlighted trimethylamine (TMA), dodecanedioic acid, citramalic acid, isobutyrylglycine, and quinaldic acid as top contributors (VIP > 2.0) (Figure 5b). Cases exhibited elevated levels of dodecanedioic acid, isobutyrylglycine, pipecolic acid, TMA, quinolinic acid, and kynurenic acid. Conversely, 1-methylhistidine, L-histidinol, methylcysteine, L-homocysteine, glycerol, and mannose-6-phosphate were reduced in cases. Additional metabolites showing significant differences between groups included 3,4-dihydroxybenzoic acid, uric acid, trans-cinnamic acid, and malonate semialdehyde (Table S1).

**Figure 5.**
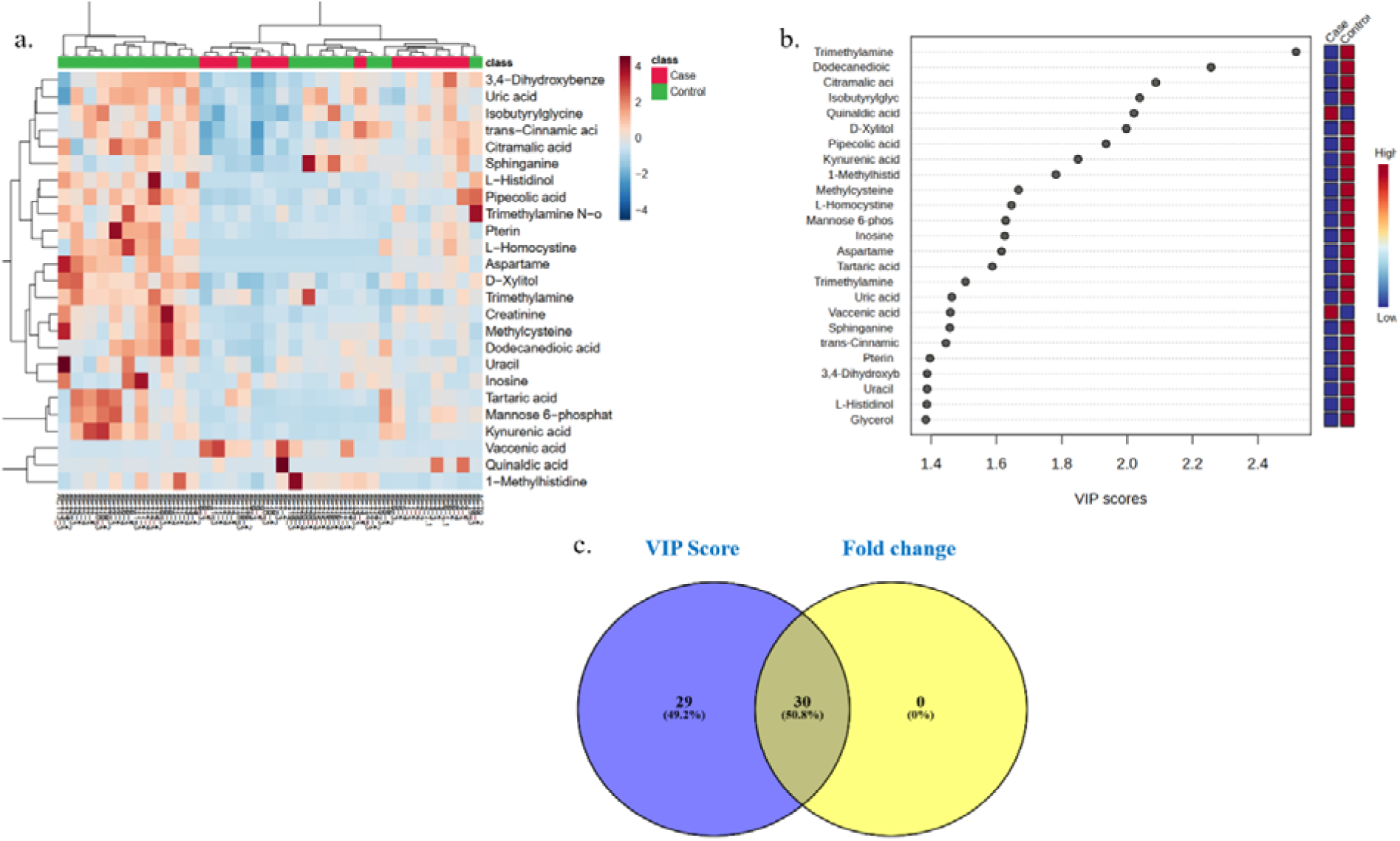
Differential serum metabolites distinguishing children with FI from controls. (a) Hierarchical clustering heatmap showing the top discriminant metabolites, with clear group separation between FI (red) and control (green) samples. (b) Variable Importance in

### Differential Metabolite Identification

Integrating statistical significance (p < 0.05) with VIP > 1.0 yielded 30 overlapping metabolites (50.8%), indicating robust discrimination (Figure 5c). Volcano plot analysis (|log□FC| > 1; adjusted p < 0.05) identified 26 downregulated and 4 upregulated metabolites in cases versus controls, consistent with deficits across amino acid metabolism, polyol biosynthesis, and lipid intermediates.

### Metabolic Pathway Enrichment Analysis

Metabolite Set Enrichment Analysis (MSEA) revealed significant perturbations in key biochemical pathways in cases compared to controls. The top enriched pathways included amino acid metabolism, serine metabolism, glycerophospholipid biosynthesis, flavin-containing monooxygenase (FMO)-mediated oxidation, and diet-dependent trimethylamine/trimethylamine N-oxide (TMA/TMAO) metabolism (Figure 6).

**Figure 6.**
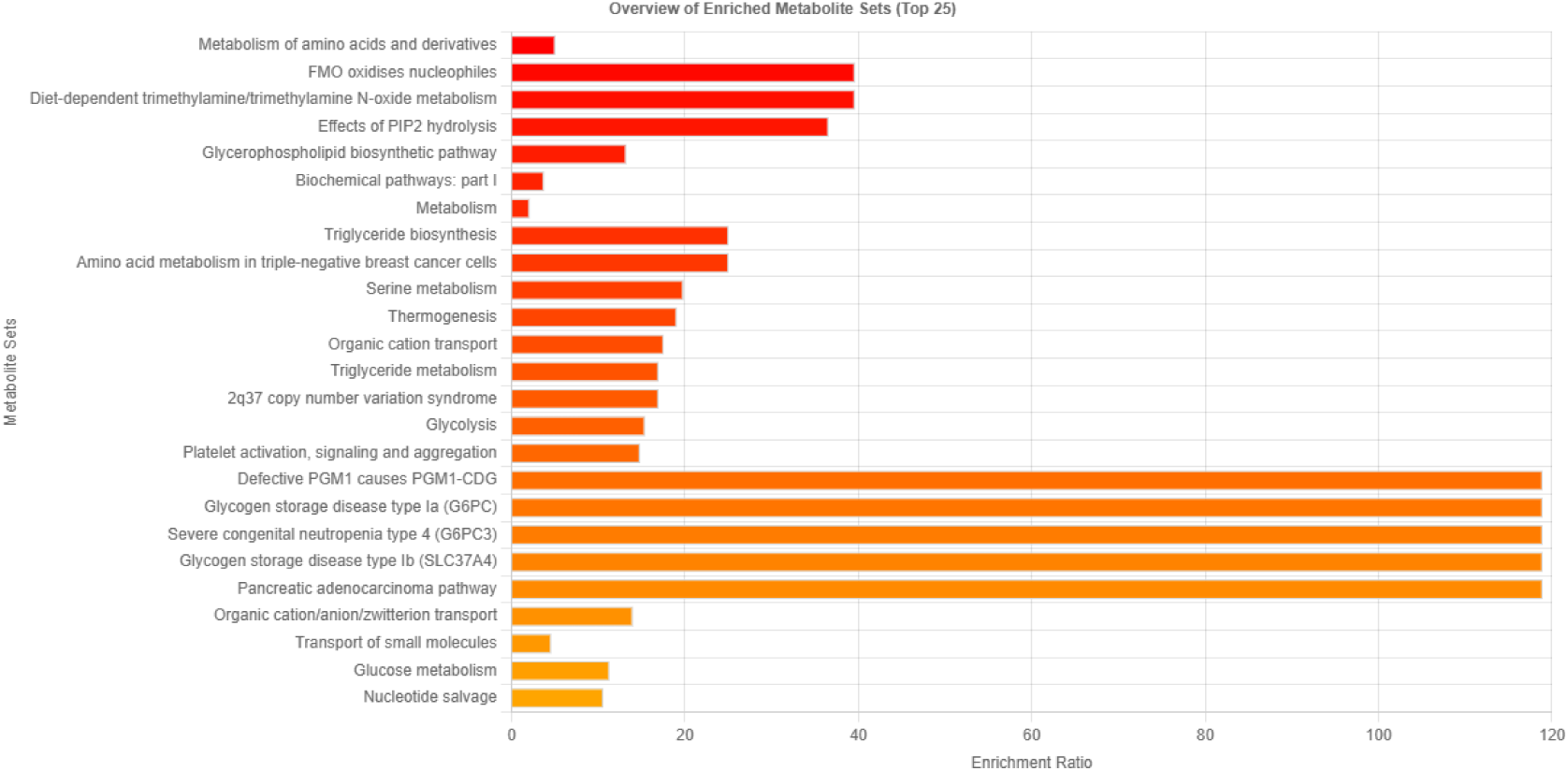
Pathway enrichment analysis of differential serum metabolites in children with FI and controls. Bar plot showing the top 25 enriched metabolite pathways ranked by enrichment ratio. Bars indicate enrichment ratios, with a colour gradient representing relative significance. Pathways related to amino acid metabolism, glycolysis, glucose metabolism, and nucleotide salvage were among the most significantly enriched in children with FI compared to controls. See Methods for details of pathway analysis and statistical thresholds.

### Disease-Based Enrichment Analysis

Disease-signature enrichment of the 30 differentially expressed metabolites (DEMs) identified 54 significant categories (FDR < 0.05) (Figure 7), with strongest overlaps for kidney disease (hits 4/28; FDR 7.72×10□□), uremia (5/92; FDR 7.72×10□□), and late-onset preeclampsia (4/40; FDR 1.74×10□□). Additional signals included early preeclampsia, colourectal cancer, chronic renal failure, Canavan disease, type 2 diabetes mellitus, and pancreatic cancer (FDR 2.13×10□□to 3.90×10□³), suggesting convergence on systemic disturbances in nitrogen, lipid, and energy metabolism consistent with the metabolic signature of cases.

**Figure 7.**
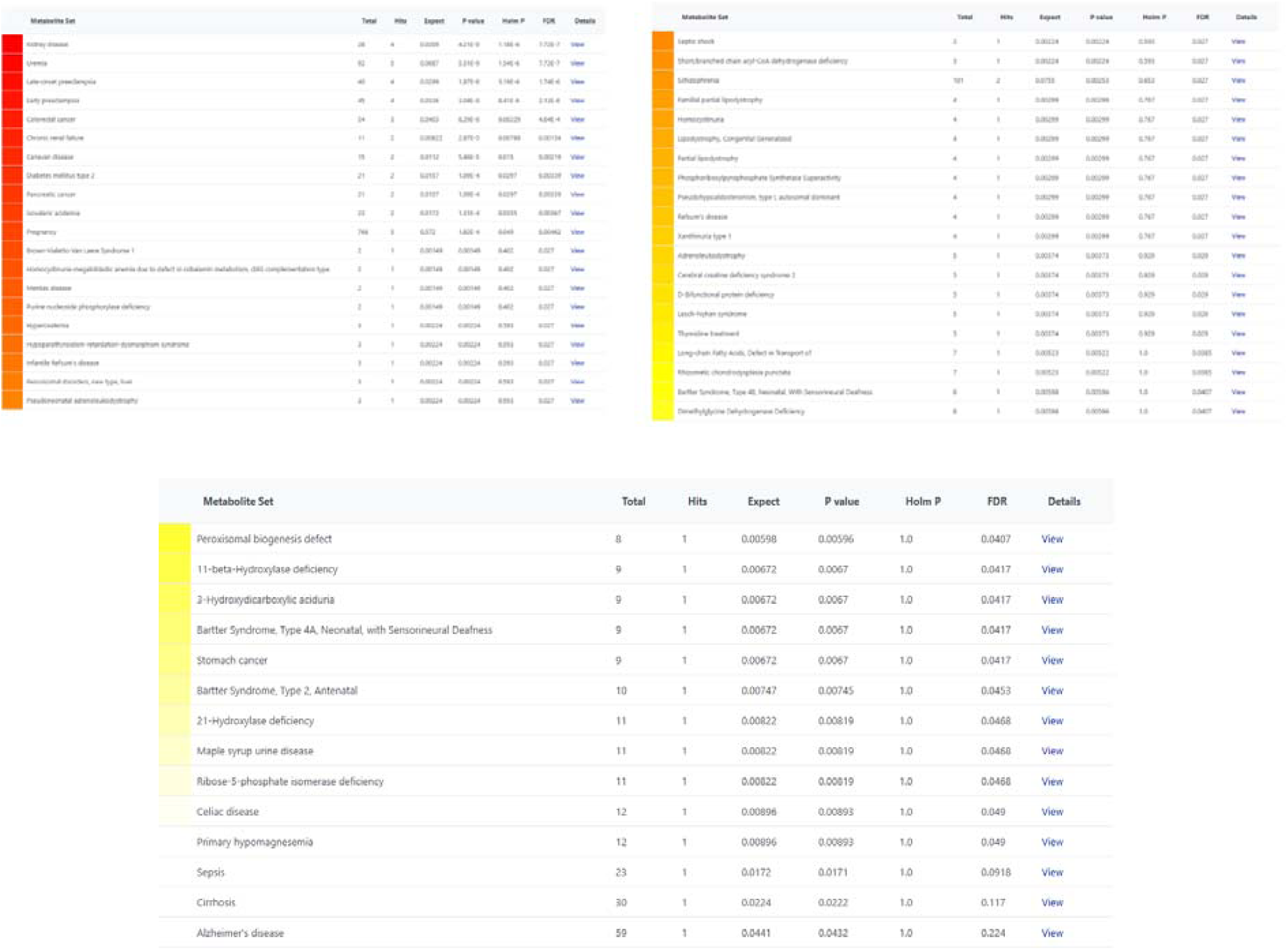
Disease enrichment analysis of differential metabolites. Table summarizing diseases significantly associated (p□<□0.05) with metabolite alterations identified in children with FI compared to controls. A total of 54 disease categories were enriched, including metabolic, inflammatory, and neurological disorders.

### Microbiome-Metabolome Integration Analysis

We profiled associations between gut genera and serum metabolites using Spearman’s correlations, revealing a structured pattern of positive and negative relationships (|r| up to ∼0.5). Fourteen correlations reached significance (*p* < 0.05) (Figure 8). Faecalibacterium—a key butyrate producer—showed the broadest connectivity, correlating positively with 1-methylhistidine (*r* = 0.493, *p* = 0.00357), 4-methoxycinnamic acid (*r* = 0.380, *p* = 0.0291), and 3,4-dihydroxyphenylglycol (r = 0.422, *p* = 0.0143), and negatively with L-homocystine (*r* = −0.352, p = 0.0447) and glycerol (*r* = −0.459, *p* = 0.00727). Clostridium exhibited five significant associations, including a positive correlation with 4-methoxycinnamic acid (*r* = 0.480, p = 0.00471) and negative correlations with D-xylitol (r = −0.379, *p* = 0.0296), glycerol (*r* = −0.371, *p* = 0.0334), 1,1-dimethylbiguanide (metformin; *r* = −0.390, *p* = 0.0247), and creatinine (r = −0.385, *p* = 0.0269). The butyrate-associated genus Butyricicoccus correlated positively with 4-methoxycinnamic acid (*r* = 0.375, *p* = 0.0317) and 3,4-dihydroxyphenylglycol (r = 0.350, *p* = 0.0457), and negatively with glycerol (*r* = −0.380, *p* = 0.0290). Roseburia showed a positive correlation with 1-methylhistidine (*r* = 0.351, *p* = 0.0453). These correlations link butyrate-producing taxa to host metabolite shifts.

**Figure 8.**
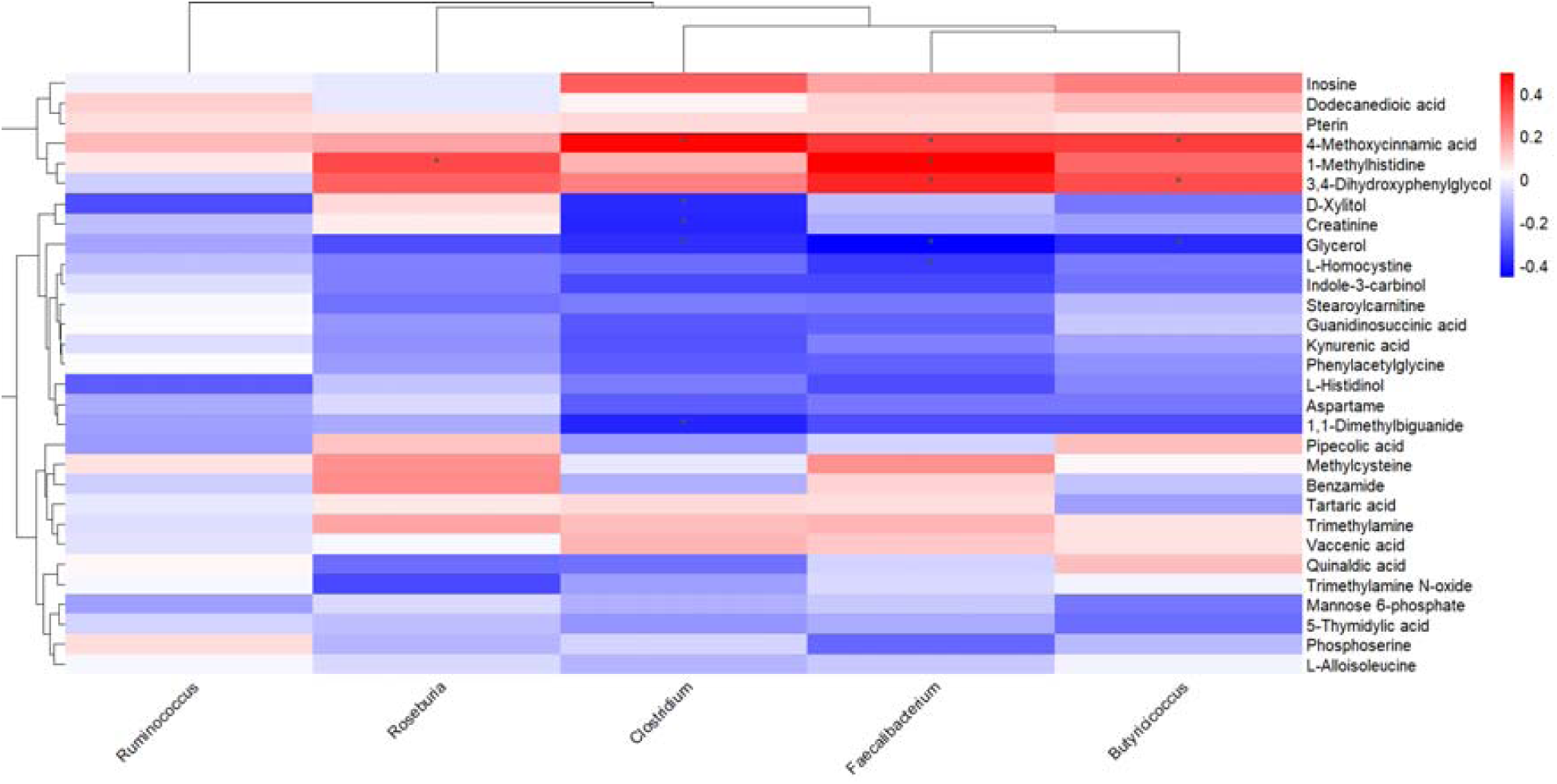
Correlation matrix between gut microbial genera and serum metabolites in children with FI and controls. Heatmap depicting significant correlations between bacterial genera (x-axis) and serum metabolites (y-axis). Spearman’s correlation coefficients were computed, and significance was adjusted using the false discovery rate (FDR). Red denotes positive and blue denotes negative correlations, with colour intensity indicating correlation strength. Asterisks mark statistically significant associations (p□<□0.05).

## Discussion

This study represents a conceptual departure in the understanding of post-surgical FI. Rather than an unavoidable sequel to surgical reconstruction, our findings indicate that FI may arise from ecological and metabolic turmoil within the gut. Multi-omics integration revealed that post-operative dysbiosis coincides with signatures of barrier disruption, impaired fatty acid metabolism, and accumulation of neuroactive and vasoactive metabolites—changes that may converge on the enteric reflex circuits governing continence (Figure 9).

**Figure 9.**
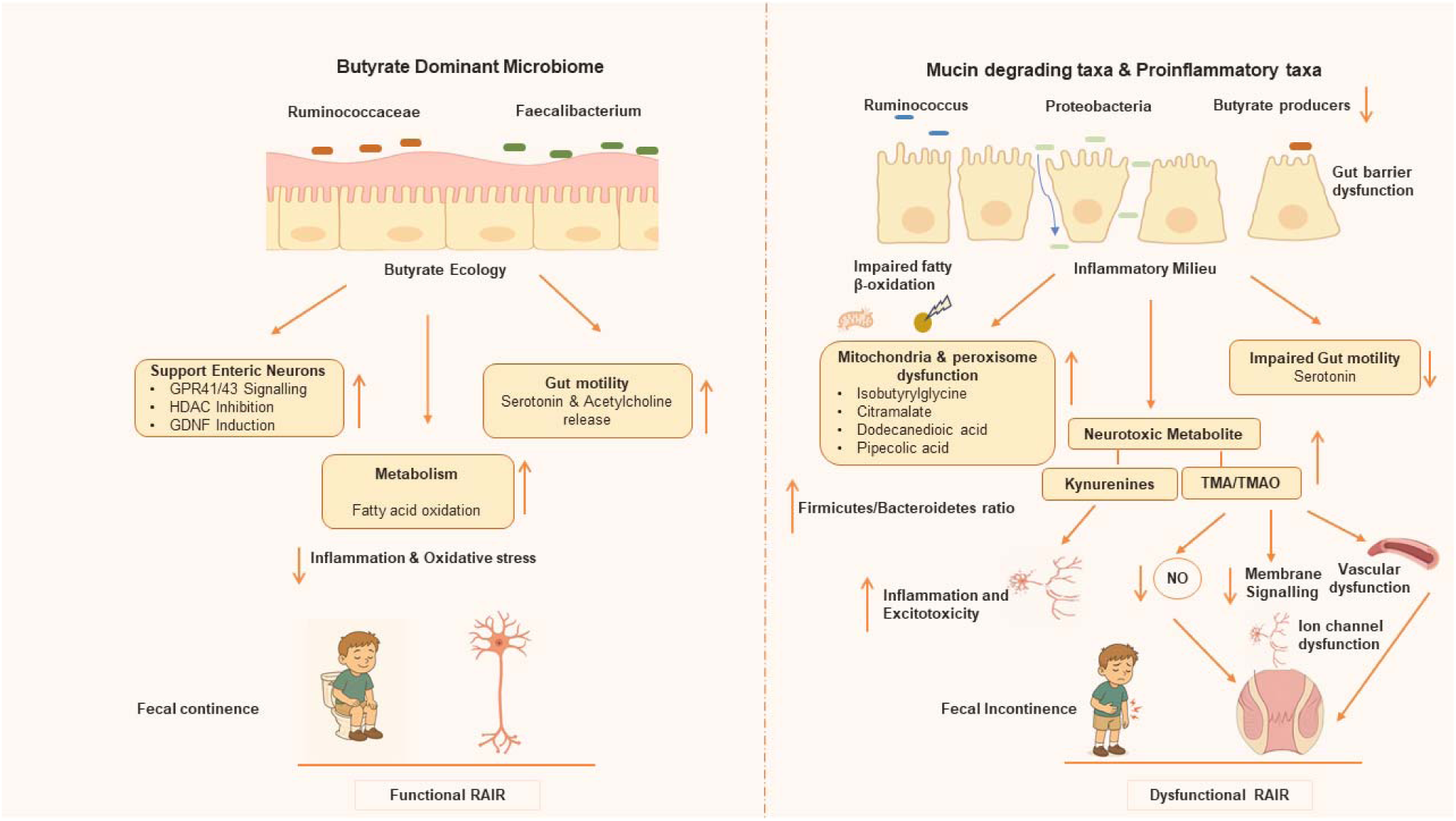
Proposed mechanistic model linking gut microbiome–metabolome interactions to continence outcomes in children with FI. Schematic representation of the contrasting ecological and metabolic states between (left) a butyrate-dominant microbiome and (right) a dysbiotic, mucin-degrading, and pro-inflammatory microbiome. In continent children, butyrate-producing taxa such as Faecalibacterium and

Our data suggest that continence is a reflection of ecological stability. Continent children, whose microbiomes were compositionally distinct by β-diversity despite comparable richness, retained Faecalibacterium and Ruminococcaceae—keystone butyrate producers that sustain gut epithelial and neuronal health^17–20^. Butyrate is a master regulator of gut homeostasis. It fuels colonocytes, strengthens tight junctions, exerts anti-inflammatory effects, and supports enteric neurons via GPR41/43 and HDAC inhibition^15,18,19,21^. It modulates ENS excitability through free fatty acid receptor 3 (FFAR3) signalling, and promotes motility through serotonin and acetylcholine release^15,18,21^ (Figure 9). It also shapes the microbial community by lowering luminal pH^22^. In our cohort, butyrate producers correlated with antioxidant metabolites and reduced redox stress, suggesting a stable host–microbe equilibrium. Experimental data corroborate this link: depletion of short-chain fatty acids (SCFAs) in rodents causes neuronal loss and dysmotility, reversible with butyrate supplementation^13^. Clinically, similar depletion is reported in functional constipation and IBS-C, paralleling our observations^23,24^.

This protective state collapses in cases. Cases exhibited an elevated Firmicutes/Bacteroidetes ratio, reflecting ecological destabilization characteristic of metabolic and inflammatory disorders^25^. We observed a shift in the microbial landscape: the depletion of butyrate-producing taxa created an ecological void, which was colonized by a hostile consortium of pathobionts. Surgical trauma, compounded by predisposing factors such as caesarean delivery, likely shifts the microbiome toward mucin-degrading taxa such as *Ruminococcus*^26^. *Ruminococcus gnavus*, for instance, erodes mucus barriers and produces pro-inflammatory polysaccharides—a pattern observed in both Crohn’s disease and systemic sclerosis, conditions that feature gut dysmotility and ENS dysfunction^27,28^. This dysbiosis favours blooms of facultative anaerobes, such as Proteobacteria (especially, Pasteurellaceae), a phylum known to thrive in inflammatory environments^29,30^. These taxa are known to produce LPS, a TLR4 ligand that has been associated with neuronal loss and dysmotility in animal models^31^. This microbial signature—the loss of guardians like *Faecalibacterium* and the rise of inflammatory opportunists—points to a shared pathway of ENS injury seen across multiple gastrointestinal disorders^28,32^ (Figure 9).

We observe that this microbial turmoil is reflected into the host’s metabolic signature. Children with FI accumulate metabolites that signal a crisis in cellular energy production. In these children, accumulation of dodecanedioic acid, a dicarboxylic acid, is associated with impaired fatty acid oxidation, together with elevated pipecolic acid, a marker of peroxisomal dysfunction, indicates organellar failure and mitochondrial overload, suggesting impaired epithelial renewal, healing, and metabolic homeostasis^33–37^. In line with this, increased levels of isobutyrylglycine are indicative of mitochondrial dysfunction and incomplete valine oxidation^38,39^. This metabolite can also signal impaired mitochondrial fatty acid β-oxidation^39,40^. This bioenergetic crisis is plausibly driven by LPS and inflammation-induced suppression of PPARα, the master regulator of fatty acid oxidation PPARα^41^. Elevated Citramalate, a microbial analogue of malate, may further disrupt TCA cycle flux and is reported in paediatric IBD, likely compounding gut barrier dysfunction.^42,43^ Strikingly, these features parallel metabolomic fingerprints in IBD and colourectal cancer, where oxidative stress, impaired peroxisome function, and mitochondrial suppression of β-oxidation underpin gut barrier dysfunction^36,44^ (Figure 9).

Beyond epithelial injury, the metabolome of children with FI is enriched with metabolites potentially toxic to the nervous system. Trimethylamine (TMA), a top discriminator, arises from microbial metabolism of choline and carnitine and is oxidized by hepatic flavin-containing monooxygenases (FMO3) to form trimethylamine-N-oxide (TMAO)^45,46^. Enriched *Roseburia* (family Lachnospiraceae; Phylum Firmicutes) and Pasteurellaceae (Gammaproteobacteria, Phylum Protoebacteria) in cases likely drive this rise^47–49^. Elevated systemic TMAO has been associated with neuroinflammation, endothelial dysfunction, and microvascular injury^46,47^. This raises the possibility that a similar toxic axis is at play between the gut and the ENS in these children. A likely physiological consequence of these disturbances is impairment of the RAIR through dysfunction of non-adrenergic, non-cholinergic (NANC) neurons that depend on nitric oxide–driven relaxation of the internal anal sphincter^50^. A TMAO-rich environment could cripple this process in two ways: directly, by interfering with nitric oxide signalling and engaging oxidative and neuroinflammatory cascades that compromise neuronal integrity^51–53^; and indirectly, by promoting endothelial dysfunction and microvascular impairment in the reconstructed anorectum^51^. This notion is strengthened by established links between vascular pathology and faecal incontinence in systemic sclerosis^28,32^. These converging insults may weaken neuromuscular support, undermine reflex integrity, and thereby explain FI in cases. In line with this, similar post-operative signatures are reported after Roux-en-Y gastric bypass, surgical hematoma evacuation, orthopaedic surgery, and coronary artery bypass grafting, where sustained TMAO elevations accompany inflammation, oxidative stress, and delayed recovery^54–57^ (Figure 9).

Lipidomic pathway enrichment offers further mechanistic clues. TMAO is known to perturb membrane lipid signalling^58^. It disrupts glycerophospholipid metabolism via MBOAT2, inducing ER stress, and activates PLC through G-protein–coupled receptors to hydrolyze PIP□^46,59–61^. Consistent with this, our pathway analysis shows significant glycerophospholipid biosynthesis enrichment and PIP□ hydrolysis. Glycerophospholipid remodelling could increase phospholipase A□ activity, with consequent production of lysophosphatidylcholine (LPC) that may amplify inflammation^62,63^. Concomitantly, sustained PIP□ hydrolysis may deplete membrane pools, limiting its availability for ion-channel regulation^64^. Such depletion could impair KCNQ-and TRPV4-channel function, destabilizing neuronal and smooth muscle excitability within the ENS^64^ ^65–67^ (Figure 9).

Inflammation also appears to redirect tryptophan metabolism. Enrichment of quinaldic acid, a kynurenine derivative, points to inflammation-driven IDO activity diverting tryptophan away from serotonin^68,69^. The result is serotonin depletion, impaired motility, and accumulation of neuroactive/neurotoxic kynurenines, which further damage ENS function^70–73^ (Figure 9). This serotonin–kynurenine axis disruption mirrors patterns in IBD, where similar metabolic shifts contribute to enteric neuronal dysfunction^69,72,74^.

The alterations of specific amino acid intermediates such as 1-methylhistidine, L-histidinol, homocysteine, and methylcysteine in FI cases is best explained by diet, as 92% of cases were vegetarian versus 50% of controls. Reduced 1-methylhistidine is a marker of low meat intake^75,76^. Decreased L-histidinol and homocysteine are consistent with limited dietary histidine and methionine from plant-based diets, although homocysteine is strongly influenced by vitamin B12 and folate status^77,78^. Methylcysteine is abundant in Allium and cruciferous vegetables, yet its levels depend on cysteine and methionine flux; with sulfur amino acid intake reduced in vegetarians, methylcysteine can also decline as seen in cases.^77,79^.

Disease enrichment analysis reveals a striking convergence between the post-surgical FI metabolome and systemic disorders. Elevated TMAO, a hallmark of our FI cohort, explains the observed overlap with chronic kidney disease, diabetes, preeclampsia and colourectal cancer^80–83^. Furthermore, the metabolic signature of FI mirrors that of diabetes and chronic kidney disease sharing key disruptions in nitric oxide and tryptophan metabolism^84–87^ (Figure 9).

Integrating these findings, we propose a dual-axis model for post-surgical FI (Figure 9). We argue that gut dysbiosis, triggered by surgery, unleashes a two-pronged pathological process: first, it induces metabolic stress that cripples cellular energy production and degrades the gut barrier; second, it disrupts host-microbe metabolism to generate a cocktail of neuroactive molecules like TMAO and kynurenines and divert tryptophan away from serotonin synthesis, a shift that impairs gut motility. These converging streams of dysfunction then inflict damage through intersecting pathways, including direct injury to the ENS and microvascular compromise that weakens the RAIR circuitry. In this model (Figure 9), the reconstructed gut, though anatomically sound, becomes an ecologically unstable environment where the neural reflexes required for continence fail.

## Limitations

These findings should be interpreted with caution. The study’s power and generalisability are constrained by a small, single-centre cohort (n=32). Its cross-sectional design precludes causal inference. Furthermore, significant group differences in diet (92% vs 50% vegetarian) and caesarean delivery rates (50% vs 15%) represent potential confounders, as both factors are known to independently shape the gut microbiome and metabolome. Methodological constraints also temper our conclusions. Although informative, 16S rRNA sequencing offered only genus-level resolution, missing the species and functional detail that shotgun metagenomics would provide. The metabolomics analysis was similarly limited; it provided a systemic snapshot from serum in positive-ion mode but did not directly probe the luminal chemistry or capture all metabolite classes. Finally, our clinical phenotyping lacked direct physiological (e.g., manometry) or biochemical (e.g., IL-6, TNF-α) data to mechanistically validate our molecular findings. Thus, the associations we report are correlational. They require rigorous validation in larger, multicentre cohorts and through direct functional experiments.

## Conclusion

This study provides new perspectives on post-surgical faecal incontinence. We redefine post-surgical faecal incontinence as a neurogastrointestinal disorder driven by gut dysbiosis rather than surgical imperfection. The results reveal associations between the host microbiome composition and post-surgical continence outcomes and identify butyrate producers as potentially important taxa linking microbial ecology to bowel function. Post-surgical FI is associated with depletion of butyrate producers and accumulation of potentially neuroactive metabolites. These changes may affect barrier function and ENS circuitry involved in continence. The parallels with IBD, IBS, and vascular disorders show that post-surgical FI extends beyond anatomy into systemic dysfunction. This insight opens the therapeutic horizon. Strategies that enrich butyrate producers, neutralize TMAO, or stabilize mitochondrial function could, in principle, reawaken dormant reflexes. Restoring continence, therefore, may depend as much on rebalancing the gut ecosystem as on refining surgical technique.

## Abbreviations

ARM: Anorectal Malformation
CKD: Chronic Kidney Disease
ENS: Enteric Nervous System
FI: Faecal Incontinence
GDNF: Glial Cell-Derived Neurotrophic Factor
HDAC: Histone Deacetylase
IBD: Inflammatory Bowel Disease
IBS-C: Irritable Bowel Syndrome with Constipation
IBS-D: Irritable Bowel Syndrome with Diarrhea
IDO: Indoleamine 2,3-Dioxygenase
IL-6: Interleukin-6
KCNQ (Kv7): Voltage-Gated Potassium Channel Subfamily Q
LPS: Lipopolysaccharide
MAMPs: Microbe-Associated Molecular Patterns
NF-κB: Nuclear Factor kappa-light-chain-enhancer of Activated B Cells
PIP□: Phosphatidylinositol 4,5-bisphosphate
RAIR: Rectoanal Inhibitory Reflex
SCFA: Short-Chain Fatty Acid
TCA cycle: Tricarboxylic Acid Cycle
TMA: Trimethylamine
TMAO: Trimethylamine N-oxide
TLR4: Toll-Like Receptor 4
TNF-α: Tumor Necrosis Factor-alpha
TRPV4: Transient Receptor Potential Vanilloid 4.

## Author Contributions

The study was conceptualized and designed by **B.P.**, who also led funding acquisition, obtained ethical approvals, performed metagenomics data analysis and interpretation, prepared graphical representations, and drafted the original manuscript, including subsequent revisions. **A.K.** contributed to data analysis and compilation of the draft, and was responsible for microbial isolation and metabolomics sample preparation. **R.J.** and **J.R.** contributed to study design, ethical approvals, supervision, and clinical data collection, and participated in manuscript review and editing. **S.S.** provided support in sample collection and processing. **A.M.** provided overall supervision and contributed to funding acquisition, methodology development, sample collection and processing, data curation, and metabolomics data analysis, in addition to reviewing and editing the manuscript.

## Data Availability Statement

The raw 16S rRNA gene sequencing data will be deposited in the NCBI Sequence Read Archive (SRA), and metabolomics data (including raw files, processed peak tables, and associated metadata) will be submitted to MetaboLights. Accession numbers will be provided upon publication.

## Ethical approval

Applicable (EC/Approval/107/2023/04/12/2023) from the Institutional Ethics Committee (IEC) of B.J. Medical College & Civil Hospital, Ahmedabad

## Informed consent

Applicable

## Acknowledgement

None

