## Supplementary Figure 1 for "Gut dysbiosis and metabolic disruption distinguish continence outcomes after anorectal malformation repair"

### Slide 1
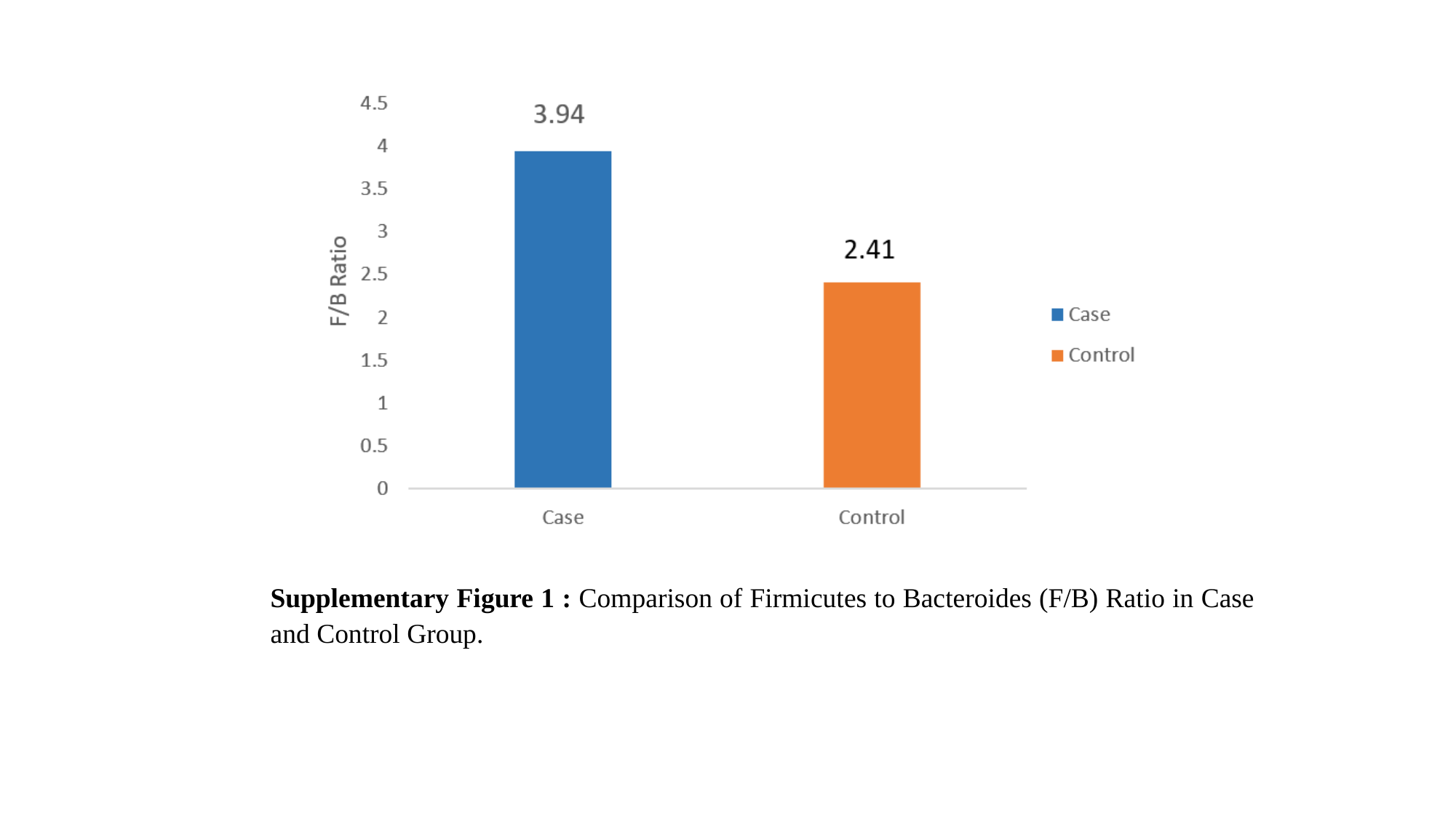

Supplementary Figure 1 : Comparison of Firmicutes to Bacteroides (F/B) Ratio in Case and Control Group.
